# Assessment of the fatality rate and transmissibility taking account of undetected cases during an unprecedented COVID-19 surge in Taiwan

**DOI:** 10.1101/2021.10.29.21265691

**Authors:** Hsiang-Yu Yuan, M. Pear Hossain, Tzai-Hung Wen, Ming-Jiuh Wang

**Author notes:** Correspondence to: Hsiang-Yu Yuan. Hsiang-Yu Yuan and M. Pear Hossain contributed equally to this article.

## Abstract

**Background:** During the COVID-19 outbreak in Taiwan between May 11 and June 20, 2021, the observed fatality rate (FR) was 5.3%, higher than the global average at 2.1%. The high number of reported deaths suggests that hospital capacity was insufficient. However, many unexplained deaths were subsequently identified as cases, indicating that there were a few undetected cases, hence resulting in a higher estimate of FR. Estimating the number of total infected cases or knowing how to reduce the undetected cases can allow an accurate estimation of the fatality rate (FR) and effective reproduction number (*R*_*t*_).

**Methods:** After adjusting for reporting delays, we estimated the number of undetected cases using reported deaths that were and were not previously detected. The daily FR and *R*_*t*_ were calculated using the number of total cases (i.e. including undetected cases). A logistic regression model was developed to predict the detection ratio among deaths using selected predictors from daily testing and tracing data.

**Results:** The estimated true daily case number at the peak of the outbreak on May 22 was 897, which was 24.3% higher than the reported number, but the difference became less than 4% on June 9 and afterward. After taking account of undetected cases, our estimated mean FR (4.7%) was still high but the daily rate showed a large decrease from 6.5% on May 19 to 2.8% on June 6. *R*_*t*_ reached a maximum value of 6.4 on May 11, compared to 6.0 estimated using the reported case number. The decreasing proportion of undetected cases was associated with the increases in the ratio of the number of tests conducted to reported cases, and the proportion of cases that are contact-traced before symptom onset.

**Conclusions:** Increasing testing capacity and tracing efficiency can lead to a reduction of hidden cases and hence improvement in epidemiological parameter estimation.

## Introduction

Knowing the actual number of coronavirus disease 2019 (COVID-19) cases throughout an outbreak is critical to provide an accurate estimate of epidemiological parameters such as the fatality rate (FR) and effective reproduction number (*R*_*t*_). These parameters aid in making proper public health decisions, assessing health care system performance, and predicting the trend of COVID-19 spread. However, the number of undetected cases can be large and may vary during an outbreak. Limited capacities for contact tracing and testing often result in underestimation of true infections ^1,2^. The proportion of undetected cases may reduce after such capacities improve. Hence, estimating this constantly changing proportion of undetected cases throughout an outbreak is important.

After several months of zero confirmed community-acquired cases, quarantine exemption for flight crews, and super spreader events in tea parlors in Wanhua in Taipei in late April and early May 2021, triggered a fresh wave of local spread of the Alpha variant ^3^. This resulted in 14,005 total reported cases between May 11 and June 20, 2021 ^4^. Approximately 5% of cases resulted in death, which was a higher case fatality rate (CFR) compared to the global rate (obtained by dividing the total number of deaths by the total number of cases worldwide), which has been consistently below 2.5% since November 16, 2020 ^5^. Whether this high CFR was mainly because of insufficient hospital capacity and treatment, or a massive proportion of undetected cases was unknown.

Early in the outbreak, testing capacity was insufficient to cope with the rising cases among initial transmission clusters. The daily number of new cases grew to more than 200 within a week and continued to increase until reaching a plateau at the end of May 2021 (i.e., 596 cases on average per day from May 22 to 28). Because of the emerging outbreak, Taiwan had been under Level 2 alert since May 11, 2021 ^6^, followed by escalation to Level 3 restrictions on May 19, 2021 ^7^, under which people are required to wear masks outdoors, gatherings of more than four people indoors and more than nine people outdoors are banned, and all schools are closed. Social distancing measures reduced individual mobility ^8^ and effectively lowered *R*_*t*_. At the same time, the daily number of tests conducted continued to increase, presumably allowing more cases to be identified.

During the outbreak, many confirmed cases failed to be detected when alive but were tested because of their death, indicating that a certain number of undetected cases existed. The number of undetected cases who eventually died (referred to as **undetected deaths**), together with the number of deaths who were known to have COVID-19 (referred to as **detected deaths**), can be used to infer the proportion of undetected cases if their fatality rates are known. Presumably, the probability of death among undetected cases is similar to that among detected cases during the early period of the outbreak when hostpial capacity and treatment is not sufficient.

Although knowing the numbers of detected and undetected deaths helps to estimate the proportion of undetected cases and hence to guide interventions, a challenge exists that many deaths from infection usually happen several weeks after symptom onset. This highlights the importance of early estimation of the true number of total cases without delay. Hence, it is important to know whether the changes in the proportion of detected deaths can be predicted by daily testing and tracing data.

We quantified time-varying FR and *R*_*t*_ by taking into account the proportion of undetected cases estimated using death data. We then developed a model based on logistic regression to predict the proportion of undetected cases using daily data related to testing and tracing capacity.

## Methods

### Data sources

We collected the date of symptom onset time and testing date for each reported death of COVID-19 from May 28 to July 22, 2021 from Taiwan Centers for Disease Control ^9^. The daily number of deaths reported before May 28 was obtained from the media. Daily number of confirmed cases was collected from Taiwan National Infectious Disease Statistics System ^4^. We collected the daily number of tests conducted from the Government Information Open Platform, Taiwan ^10,11^.

### Estimating true total cases and fatality rate

Deaths from COVID-19 were classified into two categories, detected and undetected deaths, depending on whether testing was performed before the death or not, respectively (see the schema in Figure 1A). To estimate the number of true total cases, we first considered the following ratio of undetected to detected deaths using the numbers of detected and undetected cases and their respective FR:

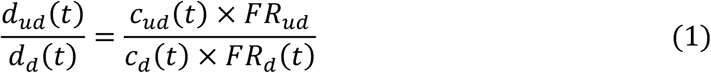

where *d*_*d*_ refers to the number of detected deaths, while *d*_*ud*_ refers to the number of undetected deaths; *c*_*d*_(*t*) and *c*_*ud*_(*t*) represent the number of cases that are detected and undetected at day *t*, respectively. Note that *t* refers to the reporting date for detected cases or detected deaths; For undetected cases or undetected deaths, *t* refers to the adjusted reporting date such that the reporting delay (i.e., the time elapsed between symptom onset and reporting) is adjusted to be the same as that of detected cases. Thus, *d*_*d*_(*t*) represents the number of deaths among the detected cases who are reported at day *t*. Similarly, *d*_*ud*_(*t*) is the number of deaths among the undetected cases whose adjusted reporting date is at day *t. FR*_*d*_(*t*), which is likely to be affected by the change in hospital capacity or treatment, represents the daily FR among the detected cases at day *t*. *FR*_*ud*_ represents the FR among the undetected cases. *FR*_*ud*_ was assumed to be a constant, estimated as the average *FR*_*d*_(*t*) during the initial two weeks (from May 11 to May 24) of the outbreak when the hostpital capacity or treatment was not sufficient. Undetected deaths who are tested later are identified as “late-detected” cases (*c*_*ld*_) (See Figure 1A). We back-projected the number of late-detected cases from their late reporting time to their adjusted reporting date *t* ^12^, using the mean and standard deviation of the reporting delay among detected cases. Our aim was to estimate *c*_*ud*_(*t*). After rearrangement, the following formula was derived:

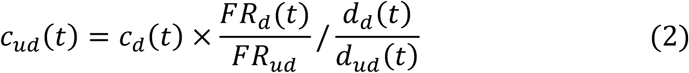

**Figure 1.**
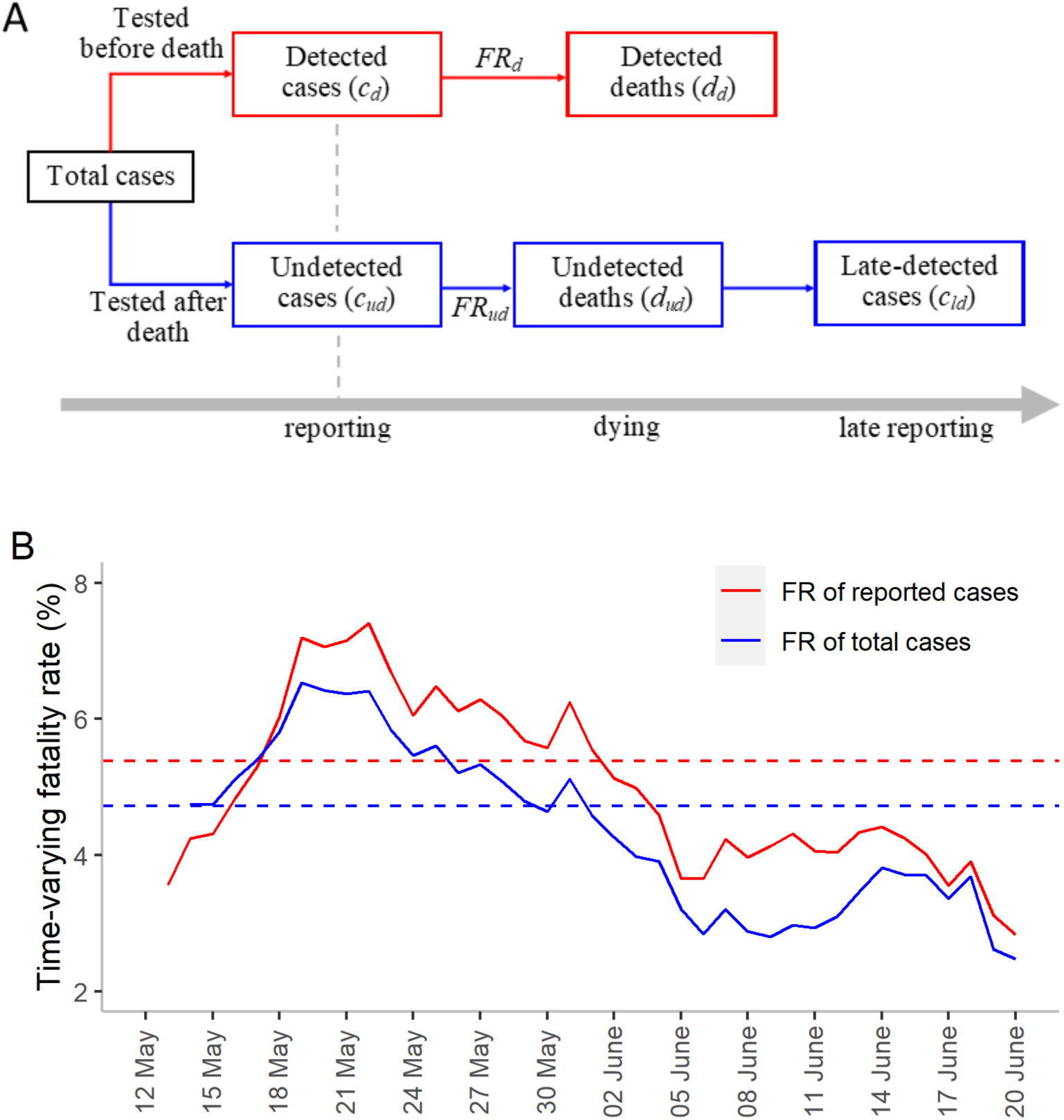
Types of cases and the fatality rate (FR). **(A)** Schema of different types of cases and deaths in relation to their testing and death time. At the time of reporting detected cases, the number of undetected cases is estimated using Eq. (2) (see Methods). *FR*_*d*_ is the FR among detected cases, and *FR*_*ud*_ is the FR among undetected cases. Reported cases include both detected and late-detected cases (after undetected deaths are tested and confirmed), while total cases include both detected and undetected cases. **(B)** Time-varying FRs among reported and total cases. The solid red line represents the proportion of reported deaths (i.e., detected and undetected deaths) among the total reported cases. The solid blue line represents the proportion of reported deaths among the total cases. The dashed red line represents the average FR among the reported cases (5.3%), whereas the blue dashed line shows the average FR among the total cases (4.7%). Note that the FR of the total cases was higher than that of the reported cases in the first few days because *FR*_*ud*_ was assumed to be same as the mean *FR*_*d*_ between May 11 and May 26. Data points during the earliest dates when the number of detected or undetected cases was zero are not shown.

The value can be solved because all of the terms on the right are either known or can be estimated. We assumed that most of the undetected deaths were identified as “late-detected” cases (*c*_*ld*_). Therefore, the number of undetected deaths was approximated by the number of late-detected cases (*d*_*ud*_ ≈ *c*_*ld*_) and then the ratio 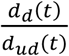 was obtained. At the same time, the proportion of detected deaths (i.e., the detection ratio among death cases; 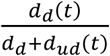 was also calculated. Finally, the true number of total cases was derived empirically as the sum of detected and undetected cases (i.e., *c*_*d*_ + *c*_*ud*_). Note that these ratios among deaths were also predicted by a regression model using data related to testing and tracing and hence a model-predicted number of total cases was obtained (see later sections).

The FRs of reported cases (including both detected and late-detected cases; *c*_*d*_ + *c*_*ld*_) and total cases were estimated at the reporting time (or the adjusted reporting time for undetected cases) using the following equations.

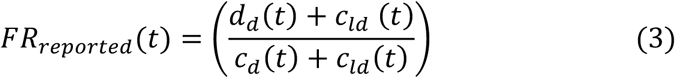

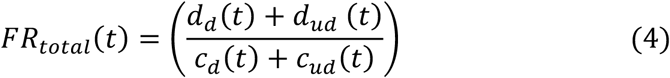

*FR*_*reported*_ is commonly known as the case fatality rate, and *FR*_*total*_ is the infection fatality rate.

### Estimating the proportion of detected deaths using a predictive model

We predicted the detection ratio among death cases using daily values of five indicators related to testing, tracing, and hospital capacities as candidate predictors. These indicators are: the ***proportion of cases without contact tracing delay, ratio of the number of tests conducted to reported cases, testing delay, reporting delay*** and ***death delay*** (for definitions, see **Error! Reference source not found**.). We calculated the delay periods in testing, reporting and death by subtracting adjusting for the date of symptom onset from the dates of these three events. Testing (the first test) earlier or on the same day as symptom onset implied that cases were contact-traced without delay. If cases were tested after symptom onset, they were either contact-traced with delay or were not contact-traced. The proportion of death cases that were contact-traced without delay was calculated.

To investigate the factors that influence the proportion of detected deaths, we developed a logistic regression model. We assumed that the number of deaths that were previously detected on day *t* follows a binomial distribution, i.e. *d*_*d*_(*t*)∼*binomial*(*d*(*t*), *m*(*t*)), where 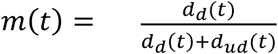 is the expected proportion of detected deaths on day *t*.

The full predictive model is:

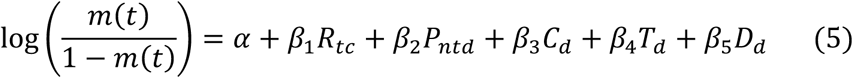

where *R*_*tc*_ is the daily ratio of tests conducted to reported cases; *P*_*ntd*_ represents the daily proportion of cases (among detected deaths) without contact tracing delay. *C*_*d*_, *T*_*d*_ and *D*_*d*_ are daily reporting, testing and death delays, respectively. *α* is the intercept and *β*_*i*_ is the regression coefficient of each covariate. The proportion of undetected COVID-19 cases can be calculated using equations (1) and (5) after *m*(*t*) is estimated:

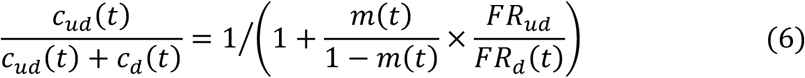

where 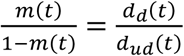 is the odds of being detected.

### Model selection

To obtain the best model, the variables in equation 5 were added to the model iteratively. First, model fit was measured for each of the variables separately using the Akaike information criterion (AIC) ^13^. The model containing the lowest AIC value was selected as the best model candidate in this batch. Next, we added one additional variable to the candidate model from the remaining four variables in the next batch. Among the two-variable models, the model with the lowest AIC value was selected as the best model candidate again. We obtained the best model candidates among three-variable, four-variable and full models. The final best model was obtained by comparing the best model candidates in different batches with the lowest AIC.

### Model validation

To evaluate whether the predictive model achieved its intended purpose (i.e., to improve the accuracy of epidemiological parameter estimation), we explored the relationship between *R*_*t*_ estimated from the total cases predicted by the best model and daily mobility data. Cases were back-projected to infection time. The result was compared with *R*_*t*_ estimated using total cases that were empirically derived or using reported cases. *R*_*t*_ estimated from four scenarios of infections were compared:

**Scenario 1 (S1): Total cases (at infection time) estimated using an empirical detection ratio –** These cases include both the reported and undetected cases at their infection time. The number of undetected cases was estimated empirically assuming reporting delay was the same for detected and undetected cases.

**Scenario 2 (S2): Total cases (at infection time) estimated from a model-predicted detection ratio –** These cases include both the reported and undetected cases at their infection time. The number of undetected cases was estimated from the model assuming reporting delay was the same for detected and undetected cases.

**Scenario 3 (S3): Reported cases (at infection time) –** Cases that were detected before death and late-detected after death at their infection time.

**Scenario 4 (S4): Reported cases (at reporting time) –** Cases that were detected before death and late-detected after death at their reporting time. Late-detected cases were back-projected at the adjusted reporting time.

### Estimating the effective reproduction number

The effective reproduction number *R*_*t*_ was estimated from the daily new cases of infection using the statistical package ***EpiEstim*** ^14^. To estimate the daily number of new cases, we assumed that both the incubation time and reporting delay followed gamma distribution ^15,16^. The mean incubation time for the circulated strain in Taiwan was 3.53 days ^17^, and we estimated the mean reporting delay as 4.45 days. Assuming the standard deviations were equal for both the distributions (estimated as 3.93 days for the reporting delay), the distribution of time between infection and reporting was gamma distribution with a mean of 7.98 days and a standard deviation of 5.28 days. The mean of the distribution was estimated as the sum of mean incubation time and confirmation delay. In contrast, the standard deviation was obtained from weighted means and pooled standard deviation for the period between infection and reporting using the following formula:

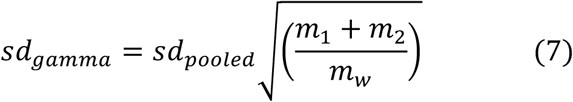

where, *m*_1_ and *m*_2_ are mean incubation time and confirmation delay and *m*_*w*_ refers weighted mean of these two. *sd*_*pooled*_ represents the pooled standard deviation for the period between infection and reporting.

We then estimated total cases at infection time using the empirical detection ratio (S1) and the model-predicted detection ratio (S2), and reported cases at infection time (S3) using a back-projection method ^12^.

We set initial conditions for estimating *R*_*t*_. Before May 11, we assumed that there were 15 cases each day between May 6 and 10, which was the average number of reported cases at infection time during this 5-day period.

## Results

Time-varying FR among true total cases (equation 4) was first quantified after taking into account undetected cases and was compared with that of reported cases. The number of total cases was also predicted using polymerase chain reaction (PCR) testing data (equations 5 and 6). To assess the impact of including undetected cases, we investigated the relationship between *R*_*t*_ generated using total cases and mobility data and then determined whether the relationship improved, compared with *R*_*t*_ from reported cases.

After the number of undetected cases was considered, the estimated FR was lower than using reported cases but was still high during the initial period of the outbreak. The mean FR of total cases was estimated to be 4.7%, which was lower than the mean FR of 5.3% for reported cases (Figure 1B). The FR increased rapidly from 4.7% and peaked at 6.5% on May 19, but then continued decreasing, reaching 2.8% on June 6. Since then, the rate was generally maintained.

From May 24 to June 3, the 5-day moving average numbers of reported cases reached a plateau and then declined thereafter (Figure 3A). The estimated true daily case number at the peak of the outbreak on May 22 was 897, which was 24.3% higher than the reported number. The difference became less than 4% on June 9 and afterward.

Until June 20, a total of 105 late-detected cases were reported, indicating many undetected deaths. Similarly, daily detected deaths also reached a plateau around May 24 (Figure 3B). However, the number of late-detected cases (at adjusted reporting time), reached a peak (7 persons per day) on May 21 and started to decline immediately, approaching zero after June 8. This indicated the improvement of the detected ratio among deaths. The detection ratio among deaths, which was about 50% initially, exceeded 95% after the end of May (Figure S1B). This ratio was very different from the observed ratio (a V-shaped pattern) without back-projection (Figure S1A).

### Predicting detection ratio using testing data

We next investigated whether the improvement in the proportion of detected cases was related to the improved capacity of testing and tracing. The indicators of the capacity were explained by the schematic of individual infection and testing statuses of each case among deaths (for definitions, please refer to Figure 2 and its legend). Depending on the time of testing, the case can be categorized as a detected death (contact-traced without delay or tested after symptom onset but before death) or an undetected death (tested after death). More efficient contact tracing allowed more cases to be traced and tested before symptom onset and was indicated by the ***proportion of cases without contact tracing delay***. This proportion fluctuated between 25% and 75% throughout the study period, with an increasing trend from late May (below 50%) to late June (above 60%) (Figure 4A). The ***testing delay*** gradually increased, from approximately two days to up to 4–6 days, until June 14, a few weeks after the outbreak started to decline (Figure 4B). The ***reporting delay*** from the day of symptom onset ranged mostly between 2.5 and 7.5 days (Figure 4E), whereas the ***death delay*** continued increasing from 5 days to more than 18 days (Figure 4C). The ***ratio of the number of tests conducted to reported cases*** increased from less than 50 to more than 200 (Figure 4D), demonstrating the improvement in testing capacity throughout the outbreak.

**Figure 2.**
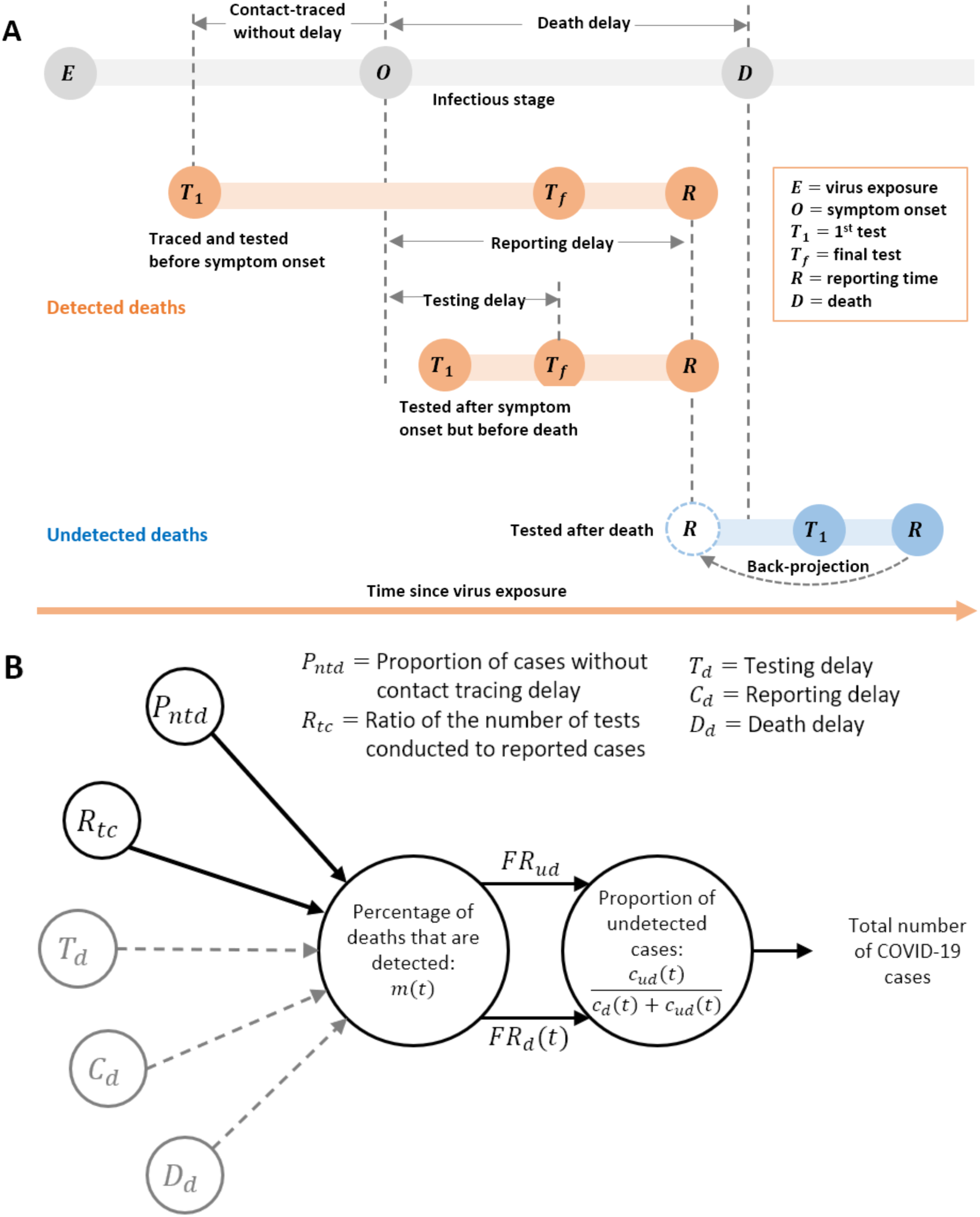
**(A)** Statuses of infection and testing of individual deaths. The gray bar represents the infection statuses of an infected case who later died after the start of infection. Orange and blue bars represent the flow of testing from the first test until the infected case is reported. The infected case was categorized as *Detected* if the first testing was performed before death. A case that was tested on the same date of or after death was categorized as *Undetected*. Among detected cases, we assumed that a case was contact traced without delay if the first test *T*_1_ was performed before symptom onset *O;* otherwise, contact traced with delay or not contact traced if the *T*_1_ was performed after symptom onset. Testing delay refers to the time between symptom onset and the last test *T*_*f*_. Similarly, the reporting delay and death delay are defined as the time difference between symptom onset and reporting, *R*, and death, *D*, respectively. The reporting time among an undetected death was adjusted to an earlier time to have the same reporting delay as detected deaths. The definitions for each status, *E, O, T*_1_, *T*_*f*_, *R* and D, are listed in the text box. **(B) Estimation of total number of COVID-19 cases (sum of detected and undetected) using a regression model**. With the best-fitting model (see Table 2), we estimated the percentage of deaths that are detected, *m*(*t*). Undetected proportion of cases was estimated based on the relationship between *m*(*t*) and fatality rates (see equation 6). Gray dashed lines represent the predictors that were not included in the best-fitting model while estimating *m*(*t*).

**Figure 3.**
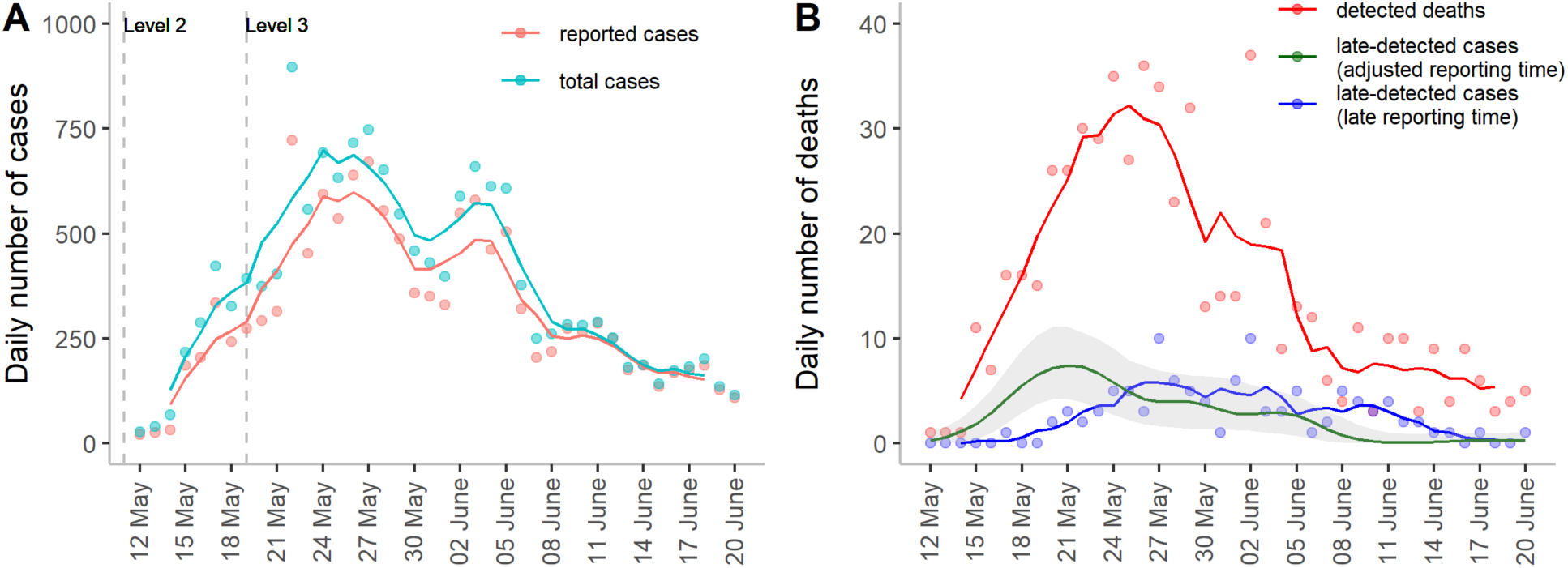
Daily numbers of reported, total cases and deaths. Data are plotted on their reporting date. **(A)** Daily number of cases that are reported. Daily number of total cases, including both the detected and undetected cases at their reporting date (green). The reporting delay of undetected cases is adjusted to be the same as that of the reported cases. The dashed vertical lines represent the implementation of level 2 and level 3 restrictions in May and June. Level 2 restrictions were started on May 11 and lasted until June 8, whereas level 3 restrictions were started on May 19 and lasted until May 28. **(B)** Daily number of deaths, plotted separately for detected deaths at their reporting time following case confirmation (red),) late-detected cases at adjusted reporting time (dark green) and late-detected cases at their late reporting time (blue). Dots represent daily numbers. Solid lines represent moving averages using a 5-day sliding window, centered at day 3 (except dark green line in (B)).

**Figure 4.**
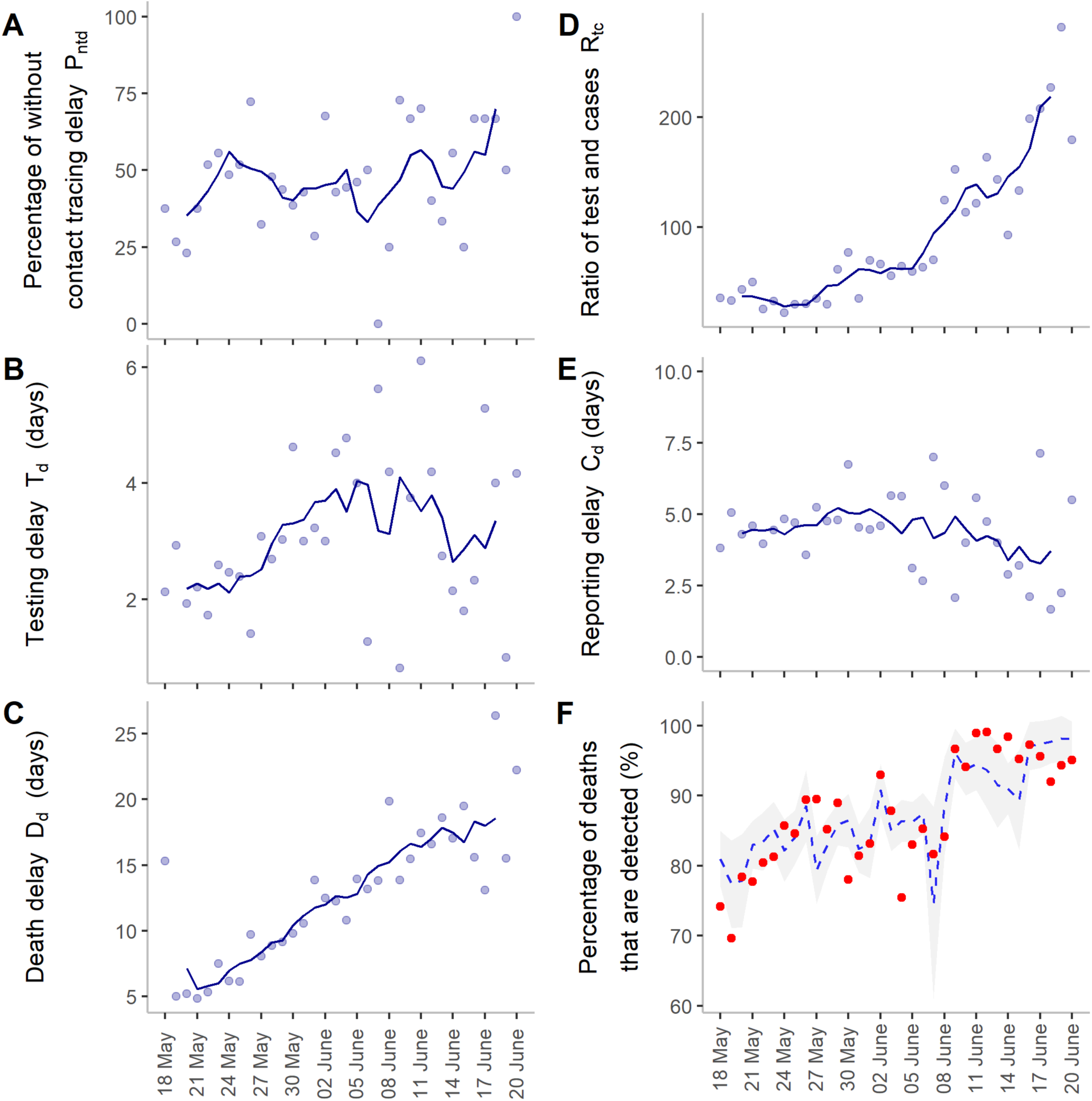
Candidate predictors that influence detected deaths. Dots in each plot represent observed values, whereas solid lines show moving averages using a 5-day sliding window, centered at day 3. **(A)** Percentage of cases without contact tracing delay was defined as the proportion of cases that were tested (the first test) earlier or on the same day as symptom onset. **(B)** Testing delay is the time delay between symptom onset and the final test. It was estimated by subtracting these two time points. **(C)** Death delay was defined as the difference between the time of death and symptom onset. **(D)** Ratio of tests to cases was calculated as the daily number of tests divided by the daily number of reported cases. **(E)** Reporting delay refers to the time delay between symptom onset and reporting. **(F)** Percentage of deaths that are detected using adjusted reported data and model prediction. Red circles represent the adjusted reported data. The blue dashed line represents the prediction results using the best fitting model. The gray shaded area represents forecasted values of the proportion of detection.

We compared models starting from the most basic to more complex ones by their AIC values to identify the best-fitting model. The model with the predictor, i.e., the proportion of cases without contact tracing delay and the ratio of tests conducted to reported cases, was selected as the best model (Model 2 in Table 1).

**Table 1.**
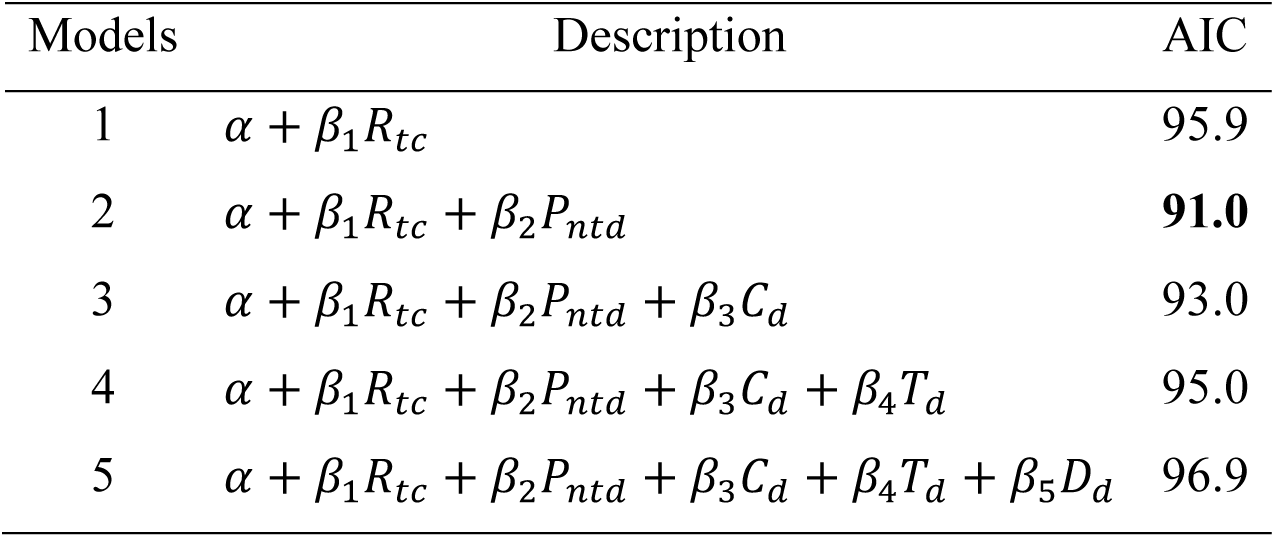
Candidate models used to choose the best model. *α* and *β*s are model coefficients, whereas the proportion of contact tracing delay (*P*_*ntd*_), the ratio of the number of tests conducted and reported cases (*R*_*tc*_), the delay in testing (*T*_*d*_), the delay in reporting (*C*_*d*_), and the delay in deaths (*D*_*d*_) are predictors. AIC represents the Akaike information criterion.

| Models | Description | AIC |
| --- | --- | --- |
| 1 | $\alpha + \beta_1 R_{tc}$ | 95.9 |
| 2 | $\alpha + \beta_1 R_{tc} + \beta_2 P_{ntd}$ | **91.0** |
| 3 | $\alpha + \beta_1 R_{tc} + \beta_2 P_{ntd} + \beta_3 C_d$ | 93.0 |
| 4 | $\alpha + \beta_1 R_{tc} + \beta_2 P_{ntd} + \beta_3 C_d + \beta_4 T_d$ | 95.0 |
| 5 | $\alpha + \beta_1 R_{tc} + \beta_2 P_{ntd} + \beta_3 C_d + \beta_4 T_d + \beta_5 D_d$ | 96.9 |

**Table 2.** Parameter estimates of the best-fitting model (Model 2)

| Covariates | Estimates | 95% Confidence intervals | p-value |
| --- | --- | --- | --- |
| Ratio of number of tests conducted to reported cases ($R_{tc}$) | 0.009 | 0.002-0.018 | 0.0180 |
| Proportion of cases without tracing delay ($P_{ntd}$) | 1.834 | 0.316-3.375 | 0.0185 |

The model successfully captured the trend in the proportion of detected deaths (Figure 4F). 20 out of 34 daily values were successfully predicted within the confidence interval. Among the values outside the interval, most of the them were in the near distance; only two dots have errors larger than two times the intervals.

The results suggest that a higher detection ratio among deaths was driven by more cases who were contact-traced without delay and a higher number of tests conducted relative to the number of cases (Table 2).

### Comparing effective reproduction number and mobility index

Comparisons were made between *R*_*t*_ estimated using i) total cases that were estimated using the empirical detection ratio; ii) total cases that were estimated from the model-predicted detection ratio using testing data; and iii) reported cases only (see Figure 5A, B, Figure S2 and Methods). When the total case number was used, *R*_*t*_ was higher during the earlier dates. The number reached a maximum value of 6.4 on May 11, compared to 6.0 estimated using the reported case number. We further evaluated the relationship between *R*_*t*_ and mobility data during the period when *R*_*t*_ reduced from the maximum value to 1 (May 11 to May 24) (Table S1). We found that when the total case number was used (either estimated using the empirical detection ratio or predicted using the testing data), a lower AIC was produced, indicating a better fit to the mobility data.

**Figure 5.**
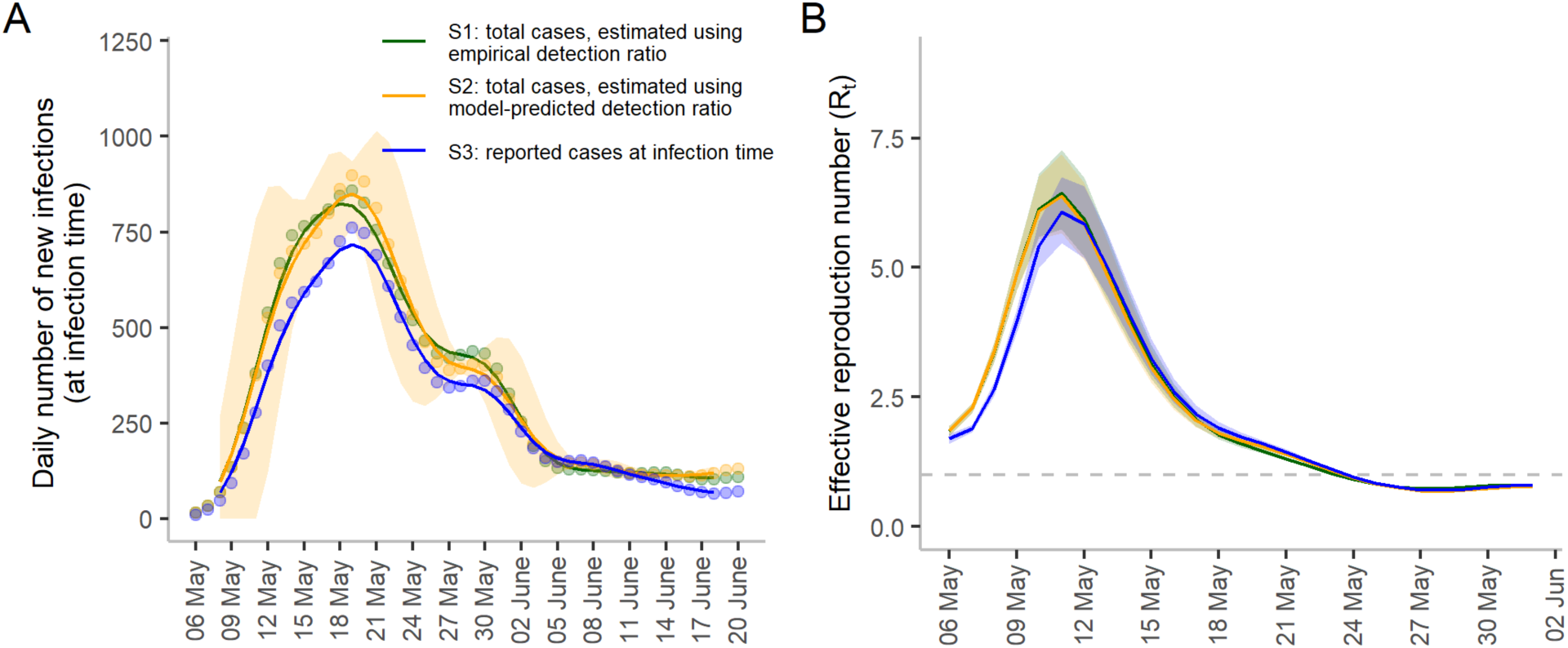
The daily number of new infections and instantaneous reproduction numbers. **(A)** The daily number of new infections was back-projected from the daily number of cases obtained from the detected and empirically estimated undetected cases (green dots; referred to as S1). The daily number of new infections obtained from the detected and model-predicted undetected cases were plotted (dark yellow dots; referred to as S2). The daily number of reported cases at their back-projected infection time (blue dots; referred to as S3). The daily numbers of new infections were back-projected from the original reported cases to virus exposure time. The lines represent moving averages using a 5-day sliding window, centered at day 3. The shaded area represents the 95% confidence interval for total cases estimated using the model-predicted detection ratio. Daily number of new detected (reported) cases at their reporting time (red dots; referred to as S4) is presented in Figure S2. **(B)** Effective reproduction number estimated from (A). Lines represent the estimated values and shaded regions represent the 95% confidence intervals. The dashed line depicts the cutoff value when *R*_*t*_ = 1. The full view of the effective reproduction number (*R*_*t*_) for the entire period between May 6 and June 20 is given in Figure S4. Color codes represent the same definition as in (A). The shaded area represents 95% confidence intervals.

In summary, efficiencies of testing and contact tracing changed during the outbreak and were useful in predicting the proportion of undetected cases. After adding the undetected cases, a better estimate of *R*_*t*_ was made and a reduction in the FR was observed.

## Discussion

Understanding whether a high FR observed in the recent largest COVID-19 outbreak in Taiwan was attributed to a higher number of undetected cases or insufficient health care capacity is important to guide interventions to reduce COVID-19 mortality in the future. An important observation is that even though the proportion of undetected cases was included, the average FR was only adjusted to 4.7% from 5.3%, which is still higher than the global average for the same time (i.e., 2.1% in May and June 2021^5^). However, the daily FR reduced to 2.8% on June 6 and remained at this low level, similar to that in the United States (i.e., 2.8% in May and June 2021 ^18^). The reduction from the initially high FR can be explained by the improvement in hospital capacity or treatment to accommodate the sudden rise in cases. This is supported by the observation that the duration between symptom onset and death among detected deaths continued increasing from approximately five days to more than two weeks in June.

The number of hidden (undetected) COVID-19 cases often affects the estimation of transmissibility of the virus and the effectiveness of non-pharmaceutical interventions (NPIs) implemented. Even though the effects of contact tracing and testing on transmissibility have been studied^19,20^, how many hidden cases do they cause is unclear. We demonstrated that the time-varying detection ratios can be predicted using data on testing and contact tracing. As a result, a more accurate *R*_*t*_ can be obtained, which is likely to be explained by mobility data better. The guidance for implementing NPIs based on changes in mobility can be provided ^8^.

We found that the ratio of the number of tests conducted to reported cases, and the proportion of cases that are contact traced without delay can be used to “nowcast” the proportion of undetected cases. Because the number of tested samples can quickly reach the capacity limit when the case number is growing, many samples remain untested. Hence, each day, the number of confirmed cases depends largely on how many tests can be performed. A day delay in testing and confirming a case, leads to a day delay in tracing the close contacts of the case. Further more, a higher contact tracing coverage together with a shorter delay of being traced enables more cases to be identified earlier^19,20^. These suggest increasing testing and tracing capacity to identify those infections earlier can reduce hidden cases more.

Modelling has been used to estimate the proportion of undetected COVID-19 cases using the observed case number during a specific period (e.g., before or after an intervention) of an outbreak^21,22^. More recently, an approach through estimating under-ascertainment by directly comparing model-predicted death with excess deaths recorded was used ^23^. We checked the number of deaths related to flu and pneumonia illness ^9^ and found no unusual excess deaths other than the reported COVID-19 deaths during this period. The proportion of undetected cases can also be calculated after incorporating seroprevalence data with false negative rates of tests into models ^24^. Overall, none of these methods estimate the constantly changing proportion of undetected cases.

Several criteria enabled us to make successful prediction using testing data. First, the number of deaths should be high. If this number is low, the uncertainty of estimating the number of undetected cases becomes high. Second, most of the deaths have to be tested eventually. Taiwan government has a strong directive to test all sudden death cases; for example, on June 18, it was announced that PCR tests would be performed for all sudden and unexplained deaths^25^. This may not likely be the case in countries with a large number of excess deaths associated with COVID-19.

In summary, predicting the number of undetected cases as early as possible using testing data can help obtain an *R*_*t*_ with a better relationship with mobility data, thus enabling policymakers to make timely public health decisions using mobility information to contain the outbreak.

## Data Availability

All data produced are available online at referenced websites.

https://data.gov.tw/dataset/120451

## Acknowledgement

The authors are indebted to City University of Hong Kong for providing excellent research facilities. The authors also acknowledge support from grants funded by Health and Medical Research Fund [COVID190215] and City University of Hong Kong [7200573 and 9610416].

## Supplementary Materials

### Establishing the relationship between mobility and effective reproduction number

Daily mobility data were obtained from Google mobility report ^26^ and were normalized after setting the mobility index on May 11 (first day of the start of the outbreak) as 1 and the value -100 as 0. The normalized mobility index ranged between 0 and 1, where higher values represent greater mobility. To compare and validate the estimated *R*_*t*_, we used a generalized linear model for Gaussian distribution with identity link function. Mobility index was adjusted in the model using the following formula adopted from a recent study ^8^:

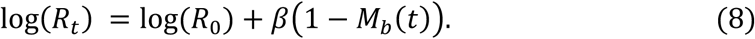

where *R*_0_ is the initial reproduction number obtained from *R*_*t*_ at the start of the outbreak (May 11, 2021), which gave the maximum number of *R*_*t*_; *M*_*b*_(*t*) represents the daily normalized mobility index; and *β* is the regression coefficient.

### Supplementary figures

**Figure S1.**
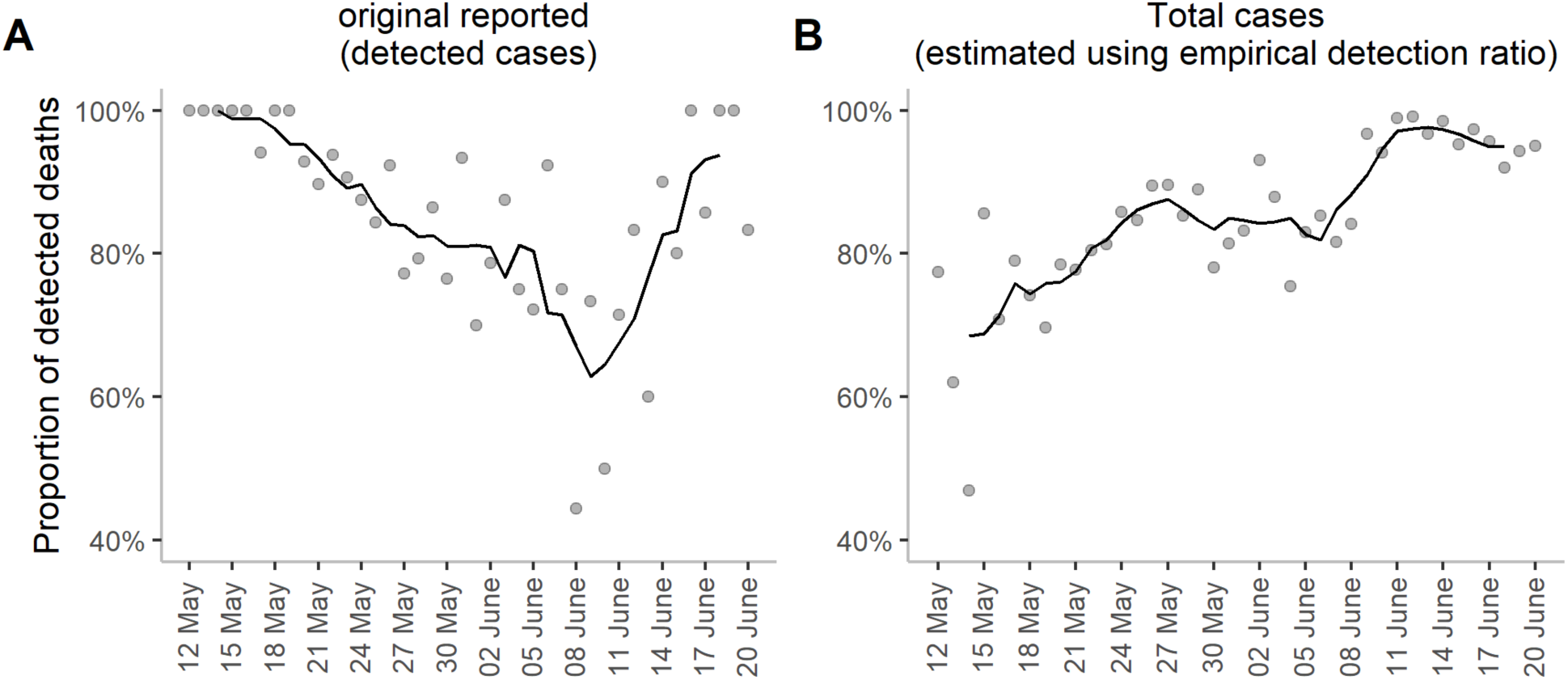
**(A)** Proportion of detected deaths among total reported deaths. **(B)** Proportion of detected deaths among total deaths estimated using the empirical detection ratio. In each plot, dots represent daily numbers that are observed or estimated. Solid lines represent moving average using a 5-day sliding window, centered at day 3.

**Figure S2.**
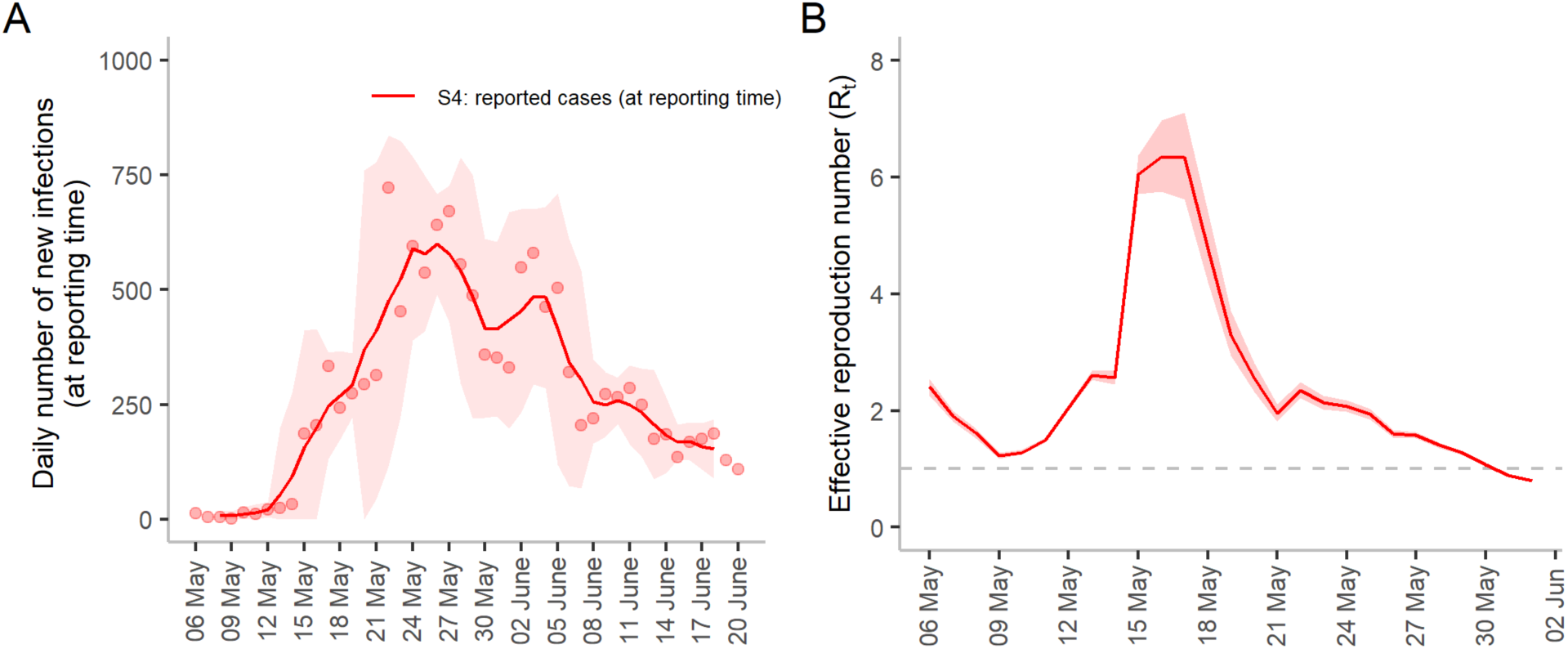
**(A)** Daily number of new infections at their reporting time. Daily values are indicated by red dots (referred to as S4 in Methods). The line represents moving averages using a 5-day sliding window, centered at day 3. **(B)** Effective reproduction number estimated from (A). The solid red line represents estimated values. The shaded area represents 95% confidence intervals. The dashed line depicts the cutoff value when *R*_*t*_ = 1. The value *R*_*t*_ during the entire period (between May 6 and June 20) is given in the Supplementary Figure S4D.

**Figure S3.**
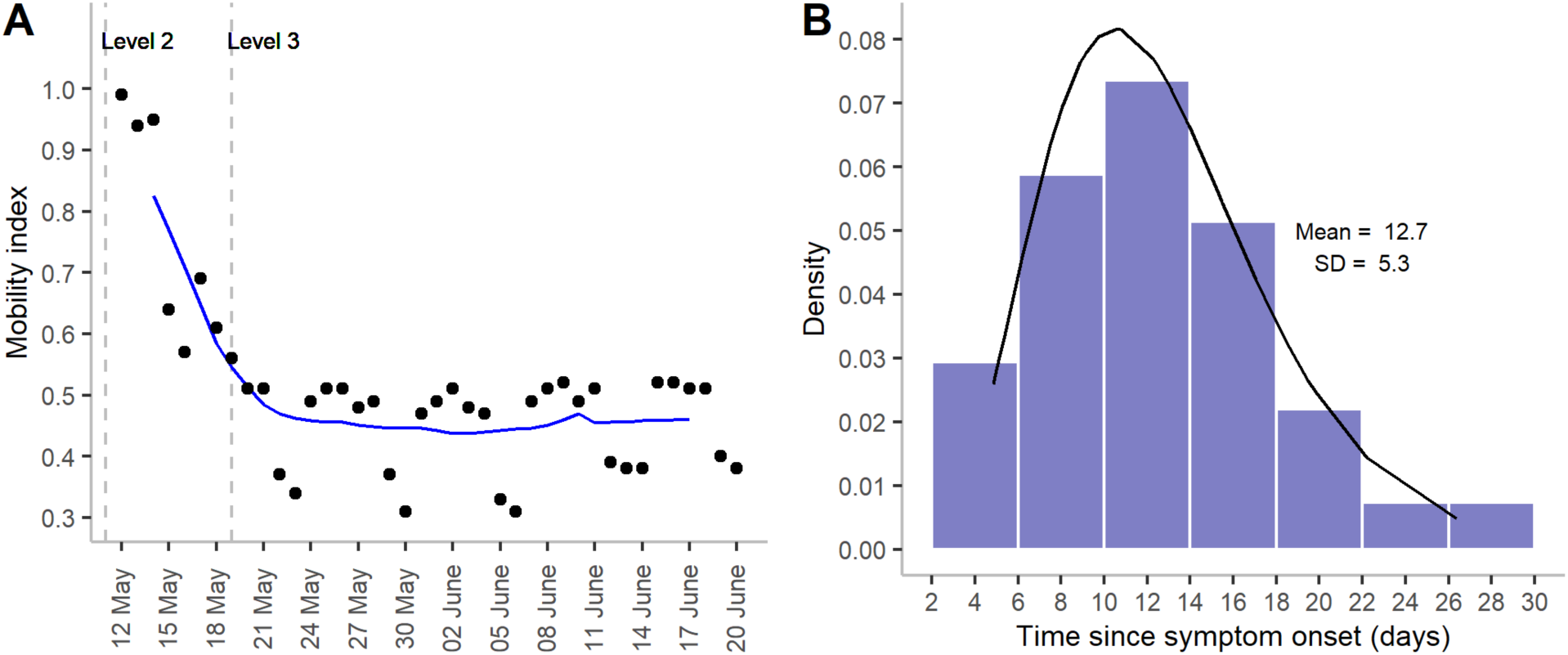
**(A)** Mobility index during the outbreak. The smooth line shows a 7-day moving average, whereas the dots represent the observed mobility index. The vertical dashed lines represent the implementation of level 2 and level 3 restrictions in May and June. Level 2 restrictions were started on May 11 and lasted until June 8, whereas level 3 restrictions were imposed for the duration between May 19 and May 28. **(B)** Distribution of death delay. The bars represent the observed frequency of delay distribution and line represents the fitted line for gamma distribution with mean and standard deviation 12.7 and 5.3 days, respectively.

**Figure S4.**
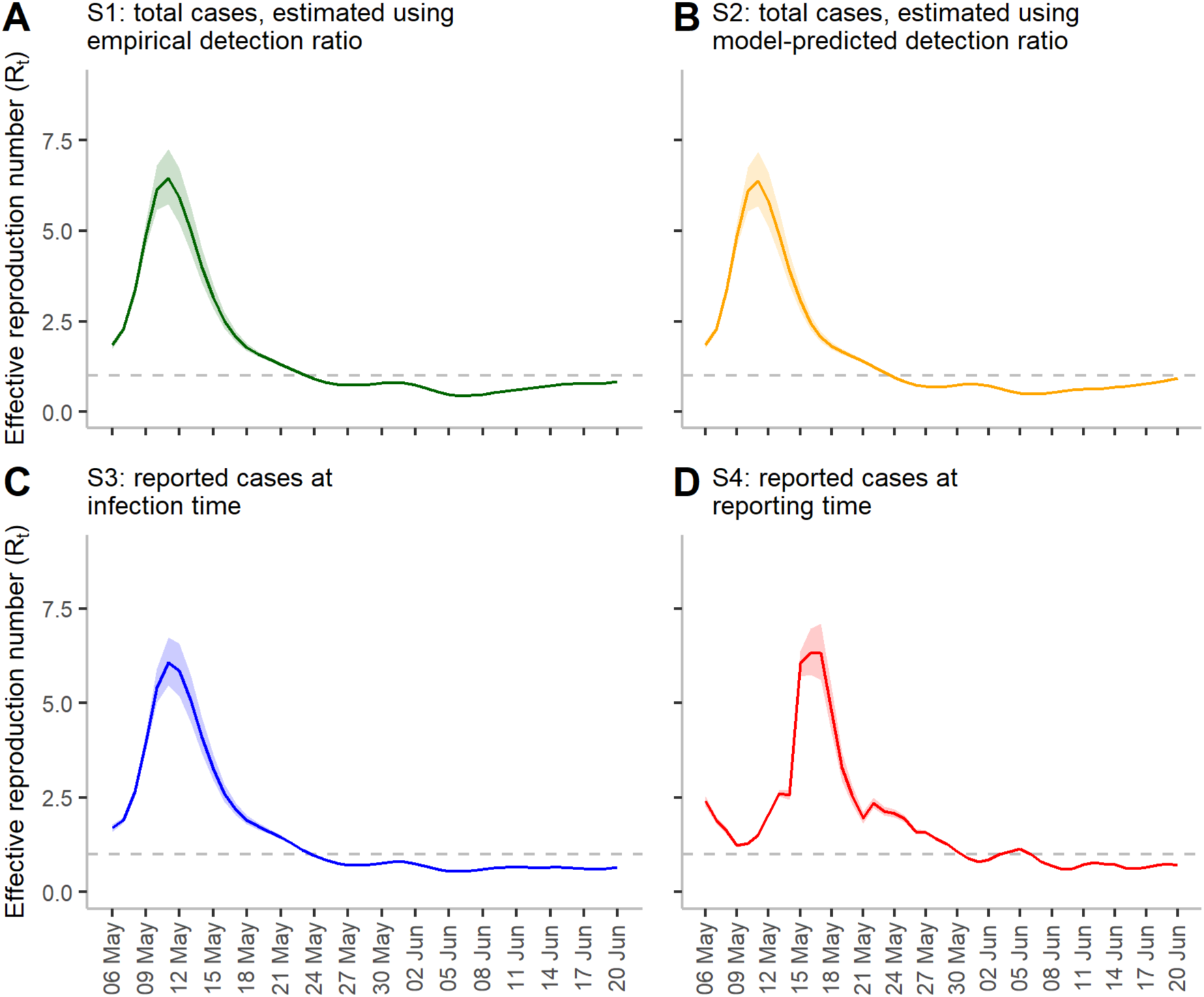
Effective reproduction number *R*_*t*_ during the entire period between May 6 and June 20. S1 and S2 refer to the numbers of total cases at infection time. S3 and S4 refer to the numbers of reported cases at infection and reporting time, respectively. Smooth solid lines represent the estimated mean *R*_*t*_, and shaded regions show the 95% confidence intervals.The dashed line depicts the cutoff value when *R*_*t*_ = 1.

### Supplementary tables

**Table S1.** Validation of the estimates of instantaneous reproduction number using mobility adjusted regression model between May 11 and May 24 when *R*_*t*_ reached one. The moving average of mobility using a 7-day sliding window, centered at day 4, was considered as the predictor. AIC represents the Akaike information criterion. Δ*AIC* shows the differences between the smallest AIC and AIC of the ith model. We rechecked the values for an extended period until May 27, when *R*_*t*_ reached a minimum. In this case, *R*_*t*_, estimated under scenario S1, showed the best fit of the mobility data with minimum AIC -27.93 (data is not presented in this table), whereas scenario S2 was treated as the second-best with AIC -27.20. The difference between the AIC of these two scenarios was less than one.

| Date | Type of data $R_t$ estimated from | Validation window<br>11 May- 24 May | |
| --- | --- | --- | --- |
| | | AIC | $\Delta AIC$ |
| At infection time | S1: Total cases estimated using the empirical detection ratio | **-27.93** | **0.00** |
|  | S2: Total cases estimated using the model-predicted detection ratio | -27.20 | 0.73 |
|  | S3: Reported cases | -21.70 | 6.23 |
| At reporting time | S4: Reported cases | 27.56 | 55.49 |

## Notes

### Competing Interest Statement

The authors have declared no competing interest.

### Author Declarations

The source data are openly available on websites listed in references.

## References

1. Grassly, N. C., Pons-Salort, M., Parker, E. P. K., White, P. J. & Ferguson, N. M. Comparison of molecular testing strategies for COVID-19 control: a mathematical modelling study. Lancet Infect. Dis. 20, (2020).

2. Contreras, S. et al. The challenges of containing SARS-CoV-2 via test-trace-and-isolate. Nat. Commun. 12, 1–13 (2021).

3. Tan, Y. COVID-19: What went wrong in Singapore and Taiwan? - BBC News. BBC News https://www.bbc.com/news/world-asia-57153195 (2021).

4. Taiwan National Infectious Disease Statistics System. Severe Pneumonia with Novel Pathogens (COVID-19). https://nidss.cdc.gov.tw/en/nndss/disease?id=19CoV (2021).

5. Our World in Data. Coronavirus (COVID-19) Deaths - Statistics and Research. https://ourworldindata.org/covid-deaths (2021).

6. Taiwan Centers for Disease Control. CECC raises epidemic warning to Level 2 and implements related restrictions and measures, effective from May 11 to June 8, in response to increased risk of community transmission. https://www.cdc.gov.tw/En/Bulletin/Detail/0jMlImCVWTuhO9mfQCd-4g?typeid=158 (2021).

7. Taiwan Centers for Disease Control. CECC raises epidemic warning to Level 3 nationwide from May 19 to May 28; strengthened measures and restrictions introduced across Taiwan to reduce community transmission. https://www.cdc.gov.tw/En/Bulletin/Detail/VN_6yeoBTKhRKoSy2d0hJQ?typeid=158 (2021).

8. Nouvellet, P. et al. Reduction in mobility and COVID-19 transmission. Nat. Commun. 12, 1–9 (2021).

9. Taiwan Centers for Disease Control. COVID-19 (SARS-CoV-2 Infection). https://www.cdc.gov.tw/En.

10. Government Information Open Platform. Taiwan’s COVID-19 coronavirus test daily delivery number. https://data.gov.tw/dataset/120451 (2021).

11. Taiwan daily confirmed local case data. https://docs.google.com/spreadsheets/d/12tQKCRuaiBZfc9yDd6tmlOdsm62ke_4AcKmNJ6q4gdU/ (2021).

12. Becker, N. G., Watson, L. F. & Carlin, J. B. A method of non-parametric back-projection and its application to aids data. Stat. Med. 10, 1527–1542 (1991).

13. Bozdogan, H. Model selection and Akaike’s Information Criterion (AIC): The general theory and its analytical extensions. Psychometrika 52, 345–370 (1987).

14. Cori, A., Ferguson, N. M., Fraser, C. & Cauchemez, S. A New Framework and Software to Estimate Time-Varying Reproduction Numbers During Epidemics. Am. J. Epidemiol. 178, 1505–1512 (2013).

15. Adam, D. C. et al. Clustering and superspreading potential of SARS-CoV-2 infections in Hong Kong. Nat. Med. 26, 1714–1719 (2020).

16. Qin, J. et al. Estimation of incubation period distribution of COVID-19 using disease onset forward time: A novel cross-sectional and forward follow-up study. Sci. Adv. 6, eabc1202 (2020).

17. Homma, Y. et al. The incubation period of the SARS-CoV-2 B1.1.7 variant is shorter than that of other strains. J. Infect. 83, e15–e17 (2021).

18. Worldometer. COVID-19 coronoavirus pandemic. https://www.worldometers.info/coronavirus/ (2021).

19. Kretzschmar, M. E. et al. Impact of delays on effectiveness of contact tracing strategies for COVID-19: a modelling study. Lancet Public Heal. 5, e452–e459 (2020).

20. Kucharski, A. J. et al. Effectiveness of isolation, testing, contact tracing, and physical distancing on reducing transmission of SARS-CoV-2 in different settings: a mathematical modelling study. Lancet Infect. Dis. 20, 1151–1160 (2020).

21. Liang, J., Yuan, H.-Y., Wu, L. & Pfeiffer, D. U. Estimating effects of intervention measures on COVID-19 outbreak in Wuhan taking account of improving diagnostic capabilities using a modelling approach. BMC Infect. Dis. 21, 1–10 (2021).

22. Li, R. et al. Substantial undocumented infection facilitates the rapid dissemination of novel coronavirus (SARS-CoV-2). Science 368, 489–493 (2020).

23. Watson, O. J. et al. Leveraging community mortality indicators to infer COVID-19 mortality and transmission dynamics in Damascus, Syria. Nat. Commun. 12, 1–10 (2021).

24. Bhattacharyya, R. et al. Incorporating false negative tests in epidemiological models for SARS-CoV-2 transmission and reconciling with seroprevalence estimates. Sci. Rep. 11, 1–14 (2021).

25. Taiwan Centers for Disease Control. Report on the Press Conference after the National Epidemic Prevention Conference on June 18-Department of Disease Control, Ministry of Health and Welfare. https://www.cdc.gov.tw/Bulletin/Detail/j5bFJQGngMHxaaQIK-j12w?typeid=9 (2021).

26. google. COVID-19 Community Mobility Reports. https://www.google.com/covid19/mobility/?hl=en-GB (2021).

